# Experiences and attitudes towards Menstrual Suppression among Women with Bacterial vaginosis Randomised to Continuous Use of NuvaRing in Kenya

**DOI:** 10.1101/2023.07.14.23292684

**Authors:** Edinah Casmir, Njeri Wairimu, Catherine Kiptinness, Lynda Oluoch, Stephen Gakuo Maina, Kristina Wilbekin Walker, Nelly Mugo, Jeanne Marrazzo, Kenneth Ngure

## Abstract

**Background:** The contraceptive vaginal ring (NuvaRing), one of the Multipurpose Prevention Technologies (MPT) products, is effective in preventing unintended pregnancies and may contribute to reducing the frequency of Bacterial Vaginosis (BV), which is a risk factor for HIV acquisition, transmission, and shedding among women. NuvaRing may cause irregular menstruation, including menstrual suppression, which may influence women’s decision on product choice, use, and acceptability. In this prospective cohort study, we assessed women’s experiences with menstruation suppression following continued NuvaRing use.

**Methods:** A total of 18 in-depth interviews were conducted using a semi-structured interview guide among purposively selected women with BV in Thika, Kenya, aged 18-40 years, who used NuvaRing continuously. All women received counseling on possibility of menstrual suppression following continuous NuvaRing use. Audio recordings were transcribed verbatim and thematically analyzed.

**Results:** Participants who expected and were aware that menstrual suppression was a possible side effect of Nuvaring accepted its benefits, and expressed acceptance and desire to continue using NuvaRing. Participants who had not anticipated menstrual suppression expressed anxiety and fear, with some expressing desire to continue using NuvaRing but worried about unintended negative consequences. The ability to perform daily activities uninterrupted, reduced expenses on sanitary supplies, enhanced sexual pleasure and relationships, and absence of menstrual pain were benefits of menstrual suppression. Although all participants received counseling on menstrual suppression, some expressed anxiety over the possibility of unintended pregnancy. As a result, they frequented medical facilities for health check-ups and pregnancy tests, and some used combined oral contraceptives to induce menses.

**Conclusion:** Understanding perceptions regarding menstrual suppression is crucial in offering targeted and comprehensive counseling to improve women’s understanding of menstruation suppression to influence acceptance and use of NuvaRing. Additionally, improved male involvement in reproductive health concerns and women’s autonomy in discussing reproductive health issues with partners is essential.

## Introduction

Women of reproductive age in sub-Saharan Africa (SSA) are faced with the double burden of risk for human immunodeficiency virus (HIV) and unintended pregnancy (1,2). Although a decline in new Human Immunodeficiency Virus (HIV) infections has been observed in this region, women account for more than half (51%) of the HIV burden (1,2) with an estimation prevalence of 6.6 % in Kenya which is twice that of men (3.1%) (3,4). This is despite the efforts to improve access to the HIV prevention technologies including pre-exposure prophylaxis (PrEP) and condoms among others (1,5). Reasons for lack and/or low uptake of these interventions could be associated with stigma, gendered differences in relationships, lack of female-initiated products, low PrEP coverage, and non-adherence with the use of the available prevention technologies. This situation calls for preventive technologies that could address the double burden and the challenges in using the products that are currently available (1,6–9).

Multipurpose Prevention Technologies (MPT) addresses the double burden of unintended pregnancy and sexually transmitted infections (STI) including HIV in reproductive age women (10). In SSA, bacterial vaginosis (BV) is found in more than 55% of women and is a substantial risk factor for HIV acquisition in, transmission to male partners from, HIV infected women, and genital HIV shedding (11,12). Pregnancy is also an independent risk factor of HIV transmission and acquisition and therefore contraception provides biomedical prevention for women with or at risk of HIV (10). The contraceptive vaginal ring (NuvaRing), a hormonal contraceptive vaginal ring containing estrogen and progesterone, recently introduced in Kenya, is effective in preventing pregnancy and may contribute to reducing frequency of BV, and HIV shedding among HIV infected women, however, it may result in irregular menstruation including menstrual suppression (6,7).

Menstruation is a normal physiological process that is surrounded by a variety of socio-cultural beliefs that may influence women’s decision-making on use of products that affect their menstrual bleeding, such as long or short duration, heavy or light, or suppression of menses (13–17). Menstruation is perceived as a process of cleansing the body by getting out the menstrual blood that is perceived to be ‘dirty blood’ (13–15,17). Literature shows that menstruation impacts on women’s daily activities, including work, school, and sexual relationships among others. Although menstruation suppression is somehow acceptable (13,18–20), literature shows that menstrual suppression is perceived to cause a build-up of “dirty” or “blocked” blood, which is in turn perceived as causing blood clots, fibroids, emotional disturbances, weight gain, infertility, or death (13,14,17,18).

A variety of social-cultural factors may influence menstrual suppression perceptions (14–16). Literature indicates little understanding and recognition of how social, cultural, interpersonal, and individual beliefs, values, and perceptions influence menstrual suppression perception and its influences on acceptability, willingness to use, and actual use of biomedical interventions (8,13–16,18,20). Understanding the end-user experiences and how they may influence acceptability and willingness to use novel prevention technologies is essential among researchers, policymakers, and service providers for informing decisions about product development and formulation, roll-out, counselling, awareness, and marketing (6–8,13,19,21). Therefore, it is crucial to understand how end-user experiences may affect acceptance and willingness to use NuvaRing, depending on its impact on hormonal changes that may ultimately cause changes in the women’s menstrual cycle, resulting in menses suppression. This paper explores how women’s experiences, perceptions and attitudes of menstrual suppression with continuous use of the NuvaRing may influence its acceptability and use in Kenyan women.

## Methods

### Study population and setting

A qualitative sub-study was conducted between May 2017 to February 2018, nested within a randomised clinical trial that aimed to determine the effects of the NuvaRing on the vaginal microbiome and BV, HIV shedding and local immunity (clinicalTrials.gov Identifier: NCT02445989). This trial, conducted at Thika Partners Study Clinic (Partners in Health Research and Development), in Central Kenya, enrolled 121 women aged 18-40 years with BV based on clinical criteria (Amsel’s criteria). Women were either HIV-infected and not on antiretroviral therapy (ART) according to Kenya ART guidelines or HIV-uninfected and, not intending to become pregnant during the study period, did not have any contraindications to hormonal contraceptive, specifically to contraceptive vaginal ring as well as capable of providing written informed consent. The participants were enrolled between April 2016 and November 2017 and randomized to continuous or cyclical NuvaRing use, with a 6-month follow-up period. However, those who accepted the alternate hormonal contraception offered at month 7 were followed up for an additional month. Women were counselled on pregnancy prevention, and, HIV prevention if HIV-uninfected; condoms were provided to the participants throughout the study. Since menstrual suppression is likely to occur among continuous users of NuvaRing, women randomised in the continuous use arm and who had used the ring for at least 2 months were interviewed to understand their experiences and attitudes towards menstrual suppression that might influence its acceptability and use among Kenyan women.

### Data collection

A total of 18 in-depth interviews (IDIs) were conducted using semi-structured interview guides aimed at identifying participants’ experiences with continuous use of NuvaRing. Topics included menstrual experiences over time, changes in menstruation before and after use of NuvaRing and participants thoughts about the changes, as well as perceptions towards menstrual suppression with NuvaRing use. Women who consented to qualitative interviews at enrolment and were willing to be interviewed to share their menstrual experiences over time with continuous use of the NuvaRing were approached to participate in the interviews. Interviews were conducted in English and Kiswahili, and audio-recorded by trained behavioural scientists in the participants’ preferred language at a private location.

### Data analysis

Audio recordings were transcribed verbatim, with Kiswahili Interviews simultaneously translated to English as needed. All transcripts were reviewed by behavioural scientists to ensure consistency and clarity, then uploaded to Dedoose Version 9.0.17 (Los Angeles, CA, USA), a web-based analysis application tool, for coding. The research team developed themes that guided development of a codebook through inductive and deductive approaches. Data were coded by two behavioural scientists after inter-coder agreement was reached. Output for specific themes (excerpts) were used to describe the study findings.

### Ethical considerations

Ethical approval was obtained from Kenya Medical Research Institute’s Scientific Review committee and Ethics Review Board (SERU). All participants gave written informed consent to participate in the study. Women and men who were unable to read or write and expressed interest in the study were offered to have the consent read to them with a witness present. All study materials were labelled with a unique identifier to prevent participants from being individually recognized by staff.

## Results

Findings from this study revealed that most participants experienced menstrual suppression with continuous use of NuvaRing. Some participants experienced suppression before and after using NuvaRing, while others experienced suppression only after NuvaRing use. A few participants experienced suppression before joining the study but did not experience it with continuous NuvaRing use, while others had never experienced suppression in their lifetime. Participants who experienced suppression before joining the study associated it to the side effects of using contraception such as depo, implants and combined oral contraceptives. Some attributed it to changes in climate, diet and stress.

Most participants reported that some women experience menstrual suppression naturally in their lifetime, while others intentionally suppressed their menses using combined oral contraception. Some women intentionally induce their menses by taking combined contraceptives, blue (dye), and drinking concentrated tea leaves. A few respondents did not know the cause of menses suppression. Notably, menstrual suppression surprised the majority of participants, despite knowing about it or having received counseling about it from providers.

Findings revealed that menstrual suppression was perceived in two different ways with most participants expressing positive attitudes, and a few expressed negative attitudes. The study findings are as summarised in the following themes, (i) perceived causes of menstrual suppression, (ii) positive attitudes towards menstrual suppression with continuous use of the NuvaRing, (iii) negative attitudes towards menstrual suppression with continuous use of the NuvaRing and (iv) male sexual partners’ perceptions on menstrual suppression.

### Perceived causes of menstrual suppression

Concerns related to menstrual suppression were not only limited to NuvaRing, because majority of the participants’ perception on major causes of menses suppression included contraception use including depot-medroxyprogesterone acetate (DMPA), emergency contraceptive pills, intrauterine device (IUD), and combined contraceptive pills. Additionally, they mentioned that stress, sickness, blood donations, changes in diet and climate can result to menses suppression. A few of the participants did not know the cause of menses suppression despite having received counseling on the possibility of menses suppression with continuous use of NuvaRing by the study staff.

> *“We can say mostly it suppresses when one starts using family planning which affects hormones. The first one is Depo, the second is this one called jadelle. Those are the one that I know mostly suppresses. Stress can cause that (suppression), sometimes change of climate” (IDI 09)*

> *“When I had exams, I had* stress *and panic and menses didn’t come the following day so I knew stress can cause that (suppression). Yes, to others, it depends with one’s hormones.” (IDI 07)*

### Positive attitudes towards menstrual suppression with continuous use of the NuvaRing

Some of the positive attitudes included ability to run daily activities uninterrupted, reduced expenses on sanitary supplies, improved sexual experience and relationships, reduced menstrual pain, blood loss, worries and odour as a result of menstruation especially those who experienced heavy menses.

Most participants who experienced menstrual suppression liked that it resulted in reduction of menstrual pain which enabled them to engage in their daily activities without pain unlike before.

> *“I feel okay that I don’t* have *the pain. I felt happy because in the past, before I got inserted the ring, I used to be moody, weak and with adnominal pain and heavy bleeding” (IDI 06)*

> *“I like staying without menses* because *you know it is stressful, maybe you are at work or any other place and then you start having backache and abdominal pain; and you don’t have restrictions in movement” (IDI 01)*

In addition, their self-esteem and comfort improved as they did not have worries and/or fears of soiling their clothes and therefore their movement was not restricted like they used to experience, before using NuvaRing, especially those who experienced heavy menses.

> *“I just feel comfortable. In the past before I joined this clinic (joining the study) I could not wear a skirt, I had to wear a trouser so that I wear a pant and hot pant to hold but now I only have a skirt and pant on and I am comfortable.” (IDI 06)*

Some participants liked menses suppression because it reduced blood loss and odour they experienced during menses before the use of NuvaRing. They said menstruation reduced blood levels, especially among people who experienced heavy menses. During menstruation, some participants had an odor that made them feel uncomfortable around people, but suppression made them to feel more comfortable and self-confident.

> *“Yes, he knows (menses suppression). He used to complain about the discharge because it used to odour, so it got to a point that the discharge ended and he was happy, he even told the doctor that the ring is good when we came here.” (IDI 09)*

Further, a few participants were satisfied with menstrual suppression, and expressed desire to purchase the NuvaRing after the study because it allowed them to save money by eliminating the need to purchase on sanitary supplies including sanitary towels, soap and water to maintain personal hygiene when on menses. They felt the much money spent on sanitary supplies would rather be spent on something else.

> *“You do not buy sanitary towels (laughing). You know in our place we buy water, so you don’t buy water often because you take a bath in the morning, afternoon and at night; you have to wash clothes and you have to buy soap. I will say that the ring is ok because I don’t feel what I used to feel in the past and it has helped me as I saved money (laughing) it is ok with me.” (IDI 06)*

> *“I am free (laughing)…from the stress of buying sanitary towels, and I save and the money for buying something else.” (IDI 07)*

Some participants experienced enhanced sexual satisfaction and relationships as their partners were happy because they could engage in sex whenever they wanted. Before suppression, some participants were uncomfortable engaging in sex during menses whenever their partners requested for it.

> *“You know when I had menses, he was not happy but now when I don’t have, he is enjoying it every day (laughing).” (IDI 01)*

A few participants were not surprised with suppression of their menses because they were aware of menstrual suppression since they had been informed by the study staff of the possibility with NuvaRing use. They also received the information before joining the study from health providers in different health facilities, as well as from peers and other women in the community. Further, some participants reported that they had experienced menstrual suppression prior to joining the study.

> *“I was told I may, or I may not get menses so I waited knowing that I may or may not get menses. I usually don’t have a problem because when you have a man in the house is when you fear.” (IDI 15)*

> *“I didn’t do a thing because before using the ring I used to use depo which suppresses menses, so I knew that.” (IDI 18)*

> *“Aah… I had not messed so I was shocked because I didn’t know what was happening, so I thought it was the ring because when you are injected (depo) you miss menses. I just waited, and when I was almost to come here but I got my menses.” (IDI 11)*

### Negative attitudes towards menstrual suppression with continuous use of the NuvaRing

Negative attitudes included possibility of unintended pregnancies, health conditions such as cervical cancer and hormonal imbalances; this prompted anxiety and fear resulting to hospital check-ups, and home-based self-pregnancy testing. Some participants expressed surprise and anxiety with menstrual suppression despite having experienced menstrual suppression before NuvaRing use and having had prior counselling/ knowledge/information on the possibility of menses suppression with the continuous use of the NuvaRing from study staff, health providers, peers and women in the community.

All participants apart from one reported the fear of possibility of unintended pregnancy and negative health consequences including infertility, hormonal imbalances, cancer, infections among others. When participants missed their menses they developed anxiety, fears and worries of the likelihood of them having conceived because they knew that they had engaged in unprotected sex. This is because they felt that sometimes contraception might fail and end up conceiving, especially given that NuvaRing was new to a few of the participants. Some of the participants reported that they wondered what could have happened because they had not engaged in sex.

> *“I thought I had* become pregnant. That is the only thing that was disturbing me. I decided that the day I come here and get told that I am pregnant I will ask them why they inserted me the ring and it does not help me and tell them to take back their thing. I really felt bad because even when I was in the house, I used to feel I had a big problem (pregnancy) ahead of me.” (IDI 16)

> *“For now, I am used to, but there is a time I used to have stress. You cannot know, for example if I am with my husband I will not tell if the family planning has worked, or it has not worked so all the time there are fears of pregnancy.” (IDI 07)*

This resulted in participants visiting the study site and other health facilities for pregnancy tests and general inquiry on the cause of suppression. Some participants said that they were afraid because they did not know what was happening with their menses and/or their health and so they decided to visit the study site and health facilities for pregnancy test and consultation. A few participants decided to buy pregnancy test kits to carry out personalized pregnancy tests. After negative pregnancy tests, both by the study staff and personalized tests, and confirmation from the study staff that they did not have any health problem, some participants reported feeling comfortable, but a few participants said they always developed anxiety whenever they experienced suppression.

> *“Aaah…. I had not messed (unprotected sex) so I was shocked because I didn’t know what was happening, so I thought it was the ring. I was not comfortable…when I came here and got explained to, I got comfortable”. (IDI 11)*

> *“Ok, I was wondering because I missed my menses the time, I expected so I wondered and asked myself like could I be pregnant and yet I have a ring? I kept quiet but I went and bought a test kit and found that I was ok, so I waited.” (IDI 04)*

> *“Mmmh…I was shocked a bit, but I said to myself that maybe they have delayed. I just went for a check-up, I asked, and I was told that they have delayed. I came here and they told me that I am ok.” (IDI 02)*

A few participants said that they did not like menstrual suppression since they did not know when to expect their menses. They reported that they had to keep their sanitary towels in their travelling bags so that they could easily access them if they received their menses unawares.

> *“I was ok but I used to plan myself well like I had sanitary towels with me always when I am on journey because it can come when I am anywhere.” (IDI 18)*

> *“I feel bad and bored because sometimes you carry sanitary towels every day and it does not come.” (IDI 01)*

These fears are informed by the community perception that if a woman does not get menses, then there is a likelihood that she is pregnant if she engages in sex, has hormonal imbalance or a health problem including infertility and that they are abnormal or bewitched and therefore they should not get married. Additionally, they said that women experience menses suppression usually develop big tummies/ bellies.

> *“…where I come from, we know that when one has menses you are not pregnant, and some say that if you don’t get menses then you can get like cancer.” (IDI 01)*

### Male sexual partners’ perceptions on menstrual suppression

The majority of the women reported that they did not disclose to their partners about their menstrual suppression because they are reserved discussing about their menses. However, 5 of the 18 women disclosed the use of NuvaRing and their menstrual suppression with their partners. One participant reported that her partner was supportive of the use of the NuvaRing because he did not like menses. A few participants said that their partners perceived menses suppression negatively because they believed it had negative effects on women’s health and therefore said that the NuvaRing was not good.

> *“He was happy because he also doesn’t like it. He knows I had planned (having contraception), and I came asked the doctor about it and I was told that missing menses is not bad because the ovaries are not there…so I explained to him so.” (IDI 01)*

> *“He tells me that it (menstrual suppression) is caused by the things I use.” (IDI 03)*

## Discussion

Findings revealed that while some women naturally experienced menstrual suppression, others intentionally suppressed their menses, and some women induced it after suppression. Most participants associated menstrual suppression with contraception use, infertility, hormonal imbalance, stress, blood donations, dietary changes, and environmental changes. Similar to reports from other studies, some participants were aware of menses suppression and its causes before and after joining the study (9,13–18,20), while others were unaware despite receiving regular menstrual suppression counseling and the possibility of suppression with continued NuvaRing use. These findings highlight participants’ different levels of knowledge/awareness on menses suppression highlighting the significance and need for continued comprehensive counselling on matters menstrual health and suppression.

Findings revealed that while some participants expressed positive attitudes concerning menstrual suppression with continued NuvaRing use, others had negative attitudes. These experiences and attitudes are worth considering because they may influence the acceptability and use of an intervention or product, in this case, NuvaRing. This finding is supported by existing literature (9,13–18,20) which show that different and varied needs, feelings, perceptions, beliefs, and knowledge/awareness based on individual, interpersonal, societal, and cultural values, norms and beliefs. In this study, participants with positive experiences expressed acceptance and a desire to continue using NuvaRing; however, those with negative attitudes had mixed emotions, with some expressing a desire to continue using NuvaRing but had fears of the unintended negative consequences. These findings have been found with the use of other products as well, such as contraception, other rings, and microbicides (13,13,18,22), showing the significance of end-user experiences in influencing the acceptability and use of an intervention or product, in this case, the NuvaRing.

Menstrual suppression was well received among participants who were aware of it, which is supported by findings from other studies (13, 9,20). Similar to these studies, participants expressed positive experiences such as ability to engage in daily activities uninterrupted, enhanced marital relationships resulting from sexual satisfaction, reduction of expenses on sanitary supplies and blood loss. Although racial and regional/ geographical differences in menses suppression exist globally, other studies have also shown that women are positive with menses suppression since it supports women’s quality of life through improving convenience, and avoiding negative menses related outcomes such as heavy and prolonged menses, nausea, bloating, fatigue and menstrual pain (13,19,20).

As other studies have shown, some participants reported negative experiences following menses suppression with a preference in non-suppression (13,18,22). Similar to these studies, we found that menstrual suppression caused worry, fear, and anxiety about unintended pregnancies, among negative health outcomes like infections, hormonal imbalances, and cancer. Existing literature demonstrate that contraceptives or ring users worry about future infertility and are uncertain about birth control effectiveness hence preventing their use or result in discontinuation (13,18,22). Although comprehensive counselling may not sufficient by itself, these concerns illustrate how crucial it is to understand the user pregnancy intentions among other concerns to support designing and providing complete counselling messages on use and adherence to avoid unintended consequences and support mental health (6,7,13–15,17,21,22).

Following pregnancy tests and a doctor’s confirmation of no underlying health issues, the majority of participants felt at ease. However, like other studies (13,16,18,19,22), a few participants had persistent fear and anxiety whenever they missed menses, and uncertainty about when they expected menses disincentivizing them from NuvaRing use. These negative experiences may be associated with existing literature (16) that the community regarded women without menses as pregnant or infertile, abnormal, bewitched, or unhealthy. As indicated in other studies, results from this study demonstrate varied participants’ knowledge of potential side effects of continued NuvaRing use on menses suppression, highlighting the importance of end-user regular tailored comprehensive counseling of an interventions use including side effects. This counseling should consider and align with individual and community levels of knowledge, needs, feelings, perceptions and concerns to address potentially harmful or negative perceptions that may influence an intervention or product acceptability, use and adherence (6,7,17,18,22).

Concerning male partners’ awareness of menses suppression, most participants felt uncomfortable discussing their menses and hence did not inform their partners about their suppression (14,15), however, a few participants did. Male partners did not like NuvaRing, except for one who disliked menses. These findings similar to other studies illustrate the major role of male influence in acceptability and use of reproductive health interventions among women, and the importance of male partner involvement in matters reproductive health to bridge the gap between acceptability and actual use (6–9,13,18). Additionally, it shows the importance of empowering women’s self-agency in discussing and disclosing matters reproductive health with their partners and male involvement in reproductive health matters to support and improve acceptability and use of new reproductive health interventions among women (6–9).

### Limitation

Studies with a large cohort and a representative sample of participants including male partners selected from various geographic locations and communities in Kenya may explain and enrich these findings since the interviews only included a small number of participants and hence may not be generalized.

## Conclusion

Exploring women’s experiences with continuous NuvaRing use in relation to menstrual changes helps develop an understanding of how menstrual suppression may influence their decision-making and preference for NuvaRing use and adherence. In this study, some participants preferred continued NuvaRing use owing to the benefits that improved women’s quality of life resulting from suppression, while others preferred non-suppression due to perceived unintended health outcomes and considered it abnormal, thus did not wish to use NuvaRing. In addition, male partners did not like NuvaRing, except for one who disliked menses. These findings are pertinent in informing the development, marketing, roll-out, education/ awareness creation, designing tailored-comprehensive counselling for users during service delivery for future MPTs product or intervention for HIV/ STIs and pregnancy prevention. Additionally, the results highlight the significance of enhancing women’s agency in discussions about reproductive health issues. Moreover, the study shows the importance of male involvement in reproductive health matters because of their crucial role in influencing reproductive health interventions or product use.

### Informed Consent

All participants provided written informed consent to participate in the study. Women and men who were unable to read or write and expressed interest in the study were offered to have the consent read to them with a witness present. All the study materials were labelled using a unique identifier.

### Ethical approval

All procedure performed in this study were in accordance with the ethical approval that was obtained from Kenya Medical Research Institute’s Scientific Review committee and Ethics Review Board (SERU).

## Data Availability

This article does not provide public access to the data described, but the corresponding author may provide it upon reasonable request at

## Acknowledgement

We thank participants who took part in the study and the study staff at the KEMRI-CCR, (PHRD)Thika Project in Kenya for all of their hard work.

## Authors Contribution

NM, JM, and KN were involved in the study’s conception, design, and funding. EC conducted the interviews, and two behavioral scientists (EC and NW) created a codebook based on the interview guide topics. EC and NW coded and reviewed the coded transcripts under KN’s supervision. EC compiled the report with manuscript review support from NW, CK, LO, SGM, KWW, NM, JM, and KN. The manuscript was submitted by EC.

